# Gaps in leprosy knowledge and hidden stigma in northwest Bangladesh: A community-based mixed-method survey among patients, contacts, communities and health workers

**DOI:** 10.64898/2026.01.20.26344485

**Authors:** Abu Sufian Chowdhury, Abhijit Saha, Johan Chandra Roy, Tanjum Ara Haque, Manoj Sarker, Rishad Choudhury Robin, Isabela De Caux Bueno, Duane Charles Hinders, Herman Wim van Brakel

## Abstract

Leprosy control requires early diagnosis, effective treatment and reduced stigma, yet community knowledge, attitudes and practices (KAP) in endemic settings remain poorly described. We assessed baseline leprosy-related KAP among key stakeholder groups in northwest Bangladesh. A community-based cross-sectional mixed-methods study was conducted in Nilphamari and Rangpur districts. We surveyed 900 respondents—index patients, close contacts, community members and health care workers—using a KAP questionnaire (score 0–9; adequate knowledge ≥7) and analyzed determinants with multivariable bootstrapped linear regression. Semi-structured interviews and focus group discussions explored explanatory models. Awareness of curability, multidrug therapy and non-contagiousness while on treatment exceeded 90% across groups, and most participants knew that disability can be prevented. Adequate knowledge was highest among health workers (89%), followed by persons affected (69%), contacts (67%) and community members (56%). Misconceptions about cause (heredity, unclean environment, supernatural or moral explanations) and transmission (skin contact, shared food) remained frequent. Higher education and being married consistently predicted better KAP, whereas older age and grade 2 disability were associated with poorer scores among index patients. Qualitative findings showed coexistence of biomedical and religious or moral explanations and highlighted internalized stigma; 42.5% of patients preferred to conceal their diagnosis despite reports of overt discrimination. In these high-endemic districts, core messages on curability and treatment have been absorbed, but knowledge gaps and hidden stigma persist among community members and vulnerable patient subgroups. Tailored, literacy-sensitive education and psychosocial support are needed to improve early detection, support post-exposure prophylaxis and reduce stigma.

## Introduction

The bacterium *Mycobacterium leprae* is the source of leprosy, a chronic infectious disease also referred to as Hansen’s disease [1]. It primarily affects the peripheral nerves and epidermis [2]. Inhaling bacilli from untreated multibacillary leprosy patients during extended close contact is considered the main mode of transmission [2,3], although the precise mechanism of transmission remains unclear [1].

Globally, about 200,000 new cases of leprosy are detected annually, with the majority from low-and middle-income countries. Brazil, India, and Indonesia account for 78.1% of new cases globally and experienced increased case detection in 2022 compared to the previous year [4]. According to the WHO, 174,087 new leprosy cases were reported in 2022, reflecting a 23.8% increase from 2021 and a prevalence rate of 0.61 per 10,000 people in South-East Asia [4].

Bangladesh remains one of the 23 WHO priority countries with a high leprosy burden and reported about 3,000 new cases in 2022 [4,5]. The country’s open border with India also poses challenges for case detection and control efforts. Early detection is vital, as untreated leprosy leads to physical disability and social consequences [6].

Knowledge and perception play important roles in how individuals recognize leprosy symptoms and seek treatment [7,8]. Adequate knowledge of leprosy and its treatment facilities is crucial for voluntary and timely reporting [9]. Perception encompasses knowledge, beliefs, attitudes, and emotions, shaped by environmental and personal factors such as culture, religion, and experience [10–12]. Lack of knowledge or misconceptions such as belief in self-cure or reliance on traditional medicine contribute to delayed diagnosis and treatment [7,13–17].

Understanding community-level knowledge, attitudes, and practices (KAP) can thus inform strategies for improved case detection and health-seeking behavior [18]. Raising awareness, reducing misconceptions, and providing accurate information are necessary to support early diagnosis and effective treatment [19].

This study is part of the international PEP++ (post-exposure prophylaxis) Project and aims to assess baseline leprosy-related knowledge, attitudes, and perceptions among index cases, close contacts, community members, and health workers in Nilphamari and Rangpur districts; high-burden regions of Bangladesh. This will help inform context-specific health communication strategies and promote behavior change in leprosy control efforts.

## Methodology

### Study design

The study used a community-based cross-sectional design. It adapted a mixed methods approach, incorporating quantitative questionnaire interviews to evaluate people’s knowledge, attitudes, and practices related to leprosy. Additionally, semi-structured interviews and Focus Group Discussions (FGD) were utilized to delve deeper into the subject matter.

### Study sites

The research took place in the northwest region of Bangladesh, specifically in Nilphamari and Rangpur districts, from October 2022 to March 2023. In 2020, the prevalence of leprosy in Nilphamari and Rangpur was recorded at 1.01 and 0.68 per 10,000 population, respectively, surpassing the national average of 0.14 per 10,000 population. These findings highlight the urgency for heightened efforts in this particular area.

### Study population and sample size calculation

The study involved four distinct groups of individuals: (1) Index Patients (those affected by Leprosy); (2) Close Contacts of Index Patients; (3) Community Members in the research areas; and (4) Health Care Workers working within the study regions.

Epi Info StatCalc for cross-sectional studies was used to calculate the quantitative sample size. For the quantitative interviews, we aimed to include a random sample of at least 100 persons of each target group in the baseline survey (index patients and close contacts). At least 170 community members have to be included. This estimate of the community members is based on an assumed prevalence of ‘negative attitudes’ of 50% at baseline and wanting to detect a reduction of 15% or more (i.e. prevalence in the 2nd survey is 35% or less). Using these parameters, a significance level of 0.05 and a power of 80%, 169 subjects are needed in each group.

In addition, we conducted interviews to gain more insight into the rationale behind people’s knowledge, beliefs attitudes and practices. We aimed to have semi-structured in-depth interviews with six persons from each participant group in each district. We also conducted one focus group discussion per participant group per district with eight to ten participants on an average. The participants for the qualitative interview were selected purposively from those who participated in the quantitative interviews; the qualitative respondents are a subset of those in the quantitative sample.

The inclusion criteria encompassed: (1) Participants who are permanent residents of Nilphamari and Rangpur districts; (2) Index patients diagnosed with leprosy in the past five years; (3) Close contacts, such as household members, family, neighbors, or social contacts, with significant interaction with the index patient (at least 20 hours per week for a minimum of three months in the year prior to the index patient’s diagnosis); (4) Community members residing in the same village or neighborhood as the index patient; and (5) Health care workers at the primary health care center within the district. Exclusion criteria consisted of individuals below 18 years of age and those unwilling or unable to provide informed consent. Close contacts, community members and health care workers were also excluded if they were or had ever been affected by leprosy. Participants who were listed as close contact of an index patient could not participate as community member also.

### Sampling method

To select index patients, first the villages were selected by stratified systematic sampling with a random start from among the villages where one or more index patient lives. A list of unions and villages in these unions where new leprosy cases were reported in the year 2016 and 2021 was prepared. Every second village from the total number of villages in each union was selected. The first index patient and village that were visited were selected randomly from the list. A total of 102 villages, spread across the 40 unions in Nilphamari and 117 villages, spread across the 44 unions in Rangpur, connected to all the 14 upazila health complexes in two districts were included.

Participants for the quantitative questionnaire interviews were selected as follows:

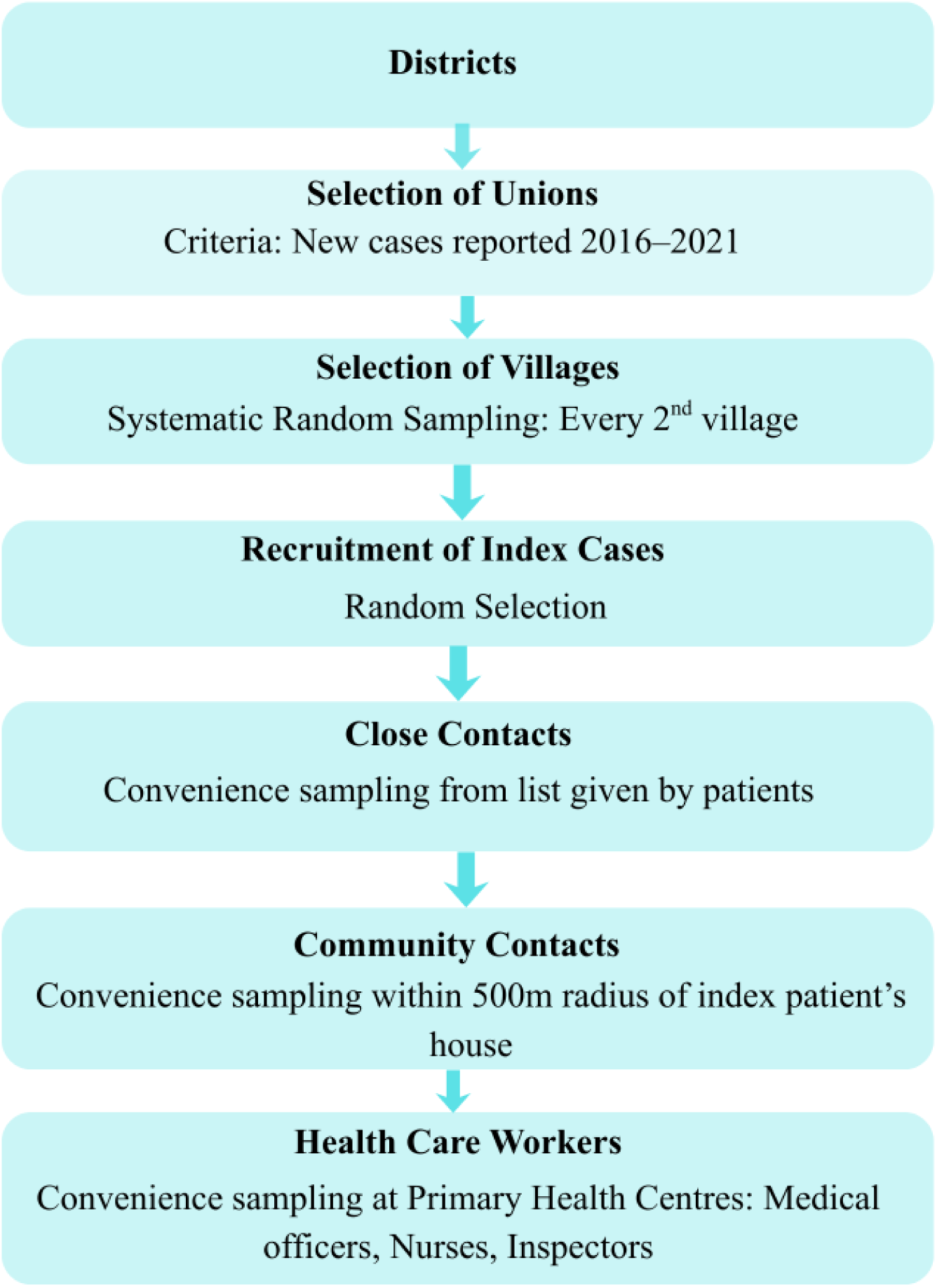

### Data collection tool

Structured questionnaire consisting of three parts was prepared for data collection. The first part of the questionnaire was related to socio-demographic characteristics of the study participants, the second part was related to assessment of participants knowledge, attitudes and practices (KAP) regarding leprosy. The KAP measure had been used in several leprosy studies between 2012 and 2017 however the results of these studies have not been published [25]. The third part (final five questions of the KAP measure), about attitudes people have towards persons affected by leprosy, were asked to index patients (*n =* 100) only. Therefore, selected psychometric properties, including floor and ceiling effects, item interpretability and internal consistency were tested and assessed based on criteria as defined by Terwee et al. 34 (26). The socio-demographic and KAP questionnaire in English and were translated into Bengali language so that it can be relevantly used in the Bangladeshi context. A back-translation was then done to English language.

Knowledge of leprosy-Based on reported response each correct response towards each item of the knowledge questionnaire; the level of knowledge towards leprosy was assessed. The questionnaire has 17 items (and consists of single and multiple answer questions). Participants could give multiple answers to some of the KAP questions. Answer options were not suggested to respondents in advance. For the questions for which multiple answers could be given, we considered an answer correct only if the correct answer was given in the absence of incorrect answers. We defined adequate knowledge as 70% or more correct answers on the knowledge section of the KAP measure (6 and above out of 9 questions). Poor knowledge was defined as 30% or less correct answers (3 and below out of 9). [25]

In addition, qualitative data on knowledge, attitudes, practices and perceptions of the participants towards leprosy were obtained using semi-structured in-depth interviews and FGD.

### Data collection procedure

Data was collected by the researchers themselves through face-to-face interview. The purpose of data collection was explained first to respondents to increase their awareness about the study before the start of the interview. They were informed regarding their voluntary participation in the study and their right to not answer any questions they did not want to. They were also ensured about regarding maintaining confidentiality of the information they provided as the researchers neither asked their name nor recorded any kind of respondent personal identity which could identify their name. A district coordinator in each district monitored the entire process.

All interviewers were trained in leprosy, in the instruments used and in the interviewing techniques prior to data collection. Pilot interviews were conducted prior to the final data collection and These participants were not included in the final sample and no changes were made to the questionnaires used. All participants were interviewed in their local language (Bengali) by a local interviewer in their home, or at a private space near their home. The in-depth interviews and FGD were audio recorded and conducted at Upazila Health Complex or Union Parishad Office.

### Data processing and analysis

The collected data were checked daily for completeness and consistency before data processing and analysis. The collected data was cleared, checked and analyzed by using tally sheet and computer. All statistical analyses were performed in two stages. In the first stage, the Knowledge, Attitude, and Practice (KAP) scale, was analyzed separately by participant type to explore group-specific distributions and patterns. In the second stage, a combined dataset including all participants was analyzed to identify overall trends and associations across variables.

Descriptive statistics were used to summarize the sociodemographic and study variables. Associations between categorical variables and outcome measures (Socio-demographic factors and KAP scores) were assessed using the Chi-square test. The Shapiro–Wilk test was applied to determine the normality of continuous variables such as age and household income. When variables did not meet the normality assumption, appropriate non-parametric tests (Mann–Whitney U or Kruskal–Wallis) were used for group comparisons.

The Spearman rank correlation test was applied to examine correlations between continuous variables and study outcomes. Variables with a *p*-value less than 0.20 in bivariate analyses were included in the multivariable models. Multiple linear regression with bootstrapped standard errors (1,000 repetitions) was employed to adjust for potential violations of normality and heteroscedasticity, and to obtain robust estimates of association.

Model validation was performed to ensure the adequacy of the regression models. The normality of residuals was examined through graphical and statistical approaches. Homoscedasticity was assessed to verify constant variance of residuals across predicted values, and multicollinearity among independent variables was evaluated using Variance Inflation Factors (VIF). A *p*-value < 0.05 was considered statistically significant. All analyses were conducted using standard statistical procedures. Nested models were compared using likelihood ratio tests to determine model improvements and best fit. All statistical analyses were performed using Stata version 18.0

### Ethics Statement

Ethical approval was obtained from the Bangladesh Medical and Research Council (BMRC) as part of a larger research project: the Post-Exposure Prophylaxis project (PEP++ project). The number of the BMRC Ethical approval is 476 28 02 2022. All participants were fully informed about the nature and objective of the study and of confidentiality of the data prior to data collection. Written consent for participation in the study was obtained from each participant in their local language. All persons approached agreed to participate in the study.

## Results

### Sociodemographic characteristics of the participants group

A total of 900 participants, of which 488 (54.2%) men were included in the study. Four groups of people were included in the study: people affected by leprosy or “index patients” (22.2%), close contacts of index patients (22.2%), community members (44.4%) and health care workers (11.1%). Most participants (89%) lived in rural areas. The average age was 39.58 years old (range 16-90); men were on average older (41.18) than women (37.69). Most people had higher (16.1%), secondary (21.4%) or primary (32.7%) education completed. Almost one third of participants were illiterate (24.9%) or could read and write but did not have any formal education (4.9%). The majority of participants was self-employed (33.6%) or had paid work (21.3%). One hundred and thirty-one (14.6%) participants were unemployed. The majority (86.9%) of participants was married. In addition, most participants were Muslim (87.6%). Only 109 participants were Hindu (12.1%). Over one third (35.9%) of the participants, excluding index patients, had a close relationship with someone with leprosy, whereas 39.1% did not and 2.8% didn’t know if they had. Index patients were on average diagnosed 11.7 months ago (range 3-28 months). Half (50%) of the health care workers who were included in this study received leprosy training. Table 1 provides an overview:

**Table 1:** Socio-demographic characteristics of participants.

|  |  | MALE |  | FEMALE |  |
| --- | --- | --- | --- | --- | --- |
|  |  | *n* | % | *n* | % |
|  | **Index patient** | 87 | 9.70 | 113 | 12.60 |
| **Participants type** | **Close contact** | 95 | 10.60 | 105 | 11.70 |
|  | **Community members** | 234 | 26.00 | 166 | 18.40 |
|  | **Health care workers** | 72 | 8.00 | 28 | 3.10 |
|  | **Total** | 488 | 54.20 | 412 | 45.80 |
| **Age Mean (Range: Male- 18-90, Female-18-75)** |  | 41.18 | - | 37.69 | - |
| **Living in rural areas** |  | 438 | 48.70 | 366 | 40.70 |
| **Knows someone with leprosy** |  | 168 | 24.00 | 155 | 22.10 |
| **Level of education completed** | **Illiterate** | 107 | 11.90 | 117 | 13.00 |
|  | **Can read and write but no formal education** | 26 | 2.90 | 18 | 2.00 |
|  | **Primary** | 149 | 16.60 | 145 | 16.10 |
|  | **Secondary** | 104 | 11.60 | 89 | 9.90 |
|  | **Higher** | 102 | 11.30 | 43 | 4.80 |
| **Occupation** | **Self-employed** | 215 | 23.90 | 87 | 9.70 |
|  | **Paid work** | 142 | 15.80 | 50 | 5.60 |
|  | **Other** | 8 | 0.90 | 83 | 9.20 |
|  | **Unemployed** | 20 | 2.20 | 111 | 12.30 |
| **Marital status** | **Married** | 409 | 45.40 | 373 | 41.40 |
|  | **Never married** | 77 | 8.60 | 17 | 1.90 |
|  | **Widowed** | 0 | 0.00 | 20 | 2.20 |
|  | **Separated** | 0 | 0.00 | 0 | 0.00 |
|  | **Divorced** | 0 | 0.00 | 1 | 0.10 |
| **Religion** | **Islam** | 431 | 47.90 | 357 | 39.70 |
|  | **Hinduism** | 56 | 6.20 | 53 | 5.90 |
|  | **Buddhism** | 1 | 0.10 | 1 | 0.10 |
|  | **Christianity** | 0 | 0.00 | 1 | 0.10 |
| **Mean (95%CI) KAP Score (best score: 9)** |  | 7.05<br>(6.88-7.22) | - | 7.11<br>(6.94-7.28) | - |

### Assessment of knowledge regarding leprosy

Recognition of key symptoms was highest for *skin patches* across all groups, particularly among health workers (93%) and persons affected (86.5%), while recognition of *loss of sensation* lagged among community members (56.25%) and contacts (63%). Misattribution of symptoms (e.g., itchiness, wounds, deformities) persisted, especially in community respondents. One of the community members mentioned during in-depth interview that

> *“Leprosy is usually caused by any skin infection or allergy that has not been treated for a long time. It is caused by allergies. As far as I know from common knowledge, it is hereditary. It can also be a consequence of [bad] actions. Allah tests people by giving them difficult diseases. This disease can happen to someone due to the religious reason…By giving some diseases, Allah takes test of a person whether that person is a believer or not, whether he/she is a good person [or a bad person]; Allah takes test of a person’s limit of patience by giving him/her a disease as He wants to see whether the person is remembering Him or forgetting Him due to the suffering from the disease” [**SSI, Community member, Male, 30-35Y”]***

Causal knowledge varied markedly: while most health workers identified *germs/bacteria* (89%), only 42% of community members did so; misconceptions (unclean environment, hereditary, supernatural/moral causes) were frequent. Transmission knowledge remained mixed, many attributed spreads to *skin contact* or *eating together*, whereas fewer correctly identified *by air*. Awareness of curability and treatment by medication was nearly universal across groups, and most respondents knew leprosy is *not contagious when on treatment* and that *disabilities can be prevented*. Overall “correct-only” performance (i.e., correct in the absence of incorrect alternatives) remained modest for cause and transmission items but was high for curability-related items. Table 2 provides an overview:

**Table 2:** An overview of the responses given per knowledge question.

|  |  |  |  |  |  |  |
| --- | --- | --- | --- | --- | --- | --- |
|  | Responses given as percentage of participants who gave the answer as *n* (%). |  |  |  |  | Percentage of people who gave the correct answer only (n = 900) |
| Early symptoms |  | Persons affected (n = 200) | Contacts (n = 200) | Community (n = 400) | Health workers (n = 100) |  |
|  | Skin patches | 173 (86.5) | 167 (83.5) | 293 (73.25) | 93 (93) | 40% |
|  | Loss of sensation | 142 (71) | 126 (63) | 225 (56.25) | 88 (88) |  |
|  | Don't know | 13 (6.5) | 19 (9.5) | 57 (14.25) | 2 (2) |  |
|  | Itchiness | 24 (12) | 21 (10.5) | 85 (21.25) | 13 (13) |  |
|  | Wounds on the skin | 11 (5.5) | 20 (10) | 51 (12.75) | 14 (14) |  |
|  | deformities or disfigurement | 20 (10) | 27 (13.5) | 67 (16.75) | 19 (19) |  |
|  | **Other: tingling, burning sensation, white patches** | 5 (2.5) | 3 (1.5) | 11 (2.75) | 4 (4) |  |
| **Cause of Leprosy** | **Don't know** | 75<br>(37.5) | 60 (30) | 145 (36.25) | 8 (8) |  |
|  | **Germs/bacteria** | 105<br>(52.5) | 115<br>(57.5) | 168 (42) | 89 (89) | 38% |
|  | **Unclean environment** | 33<br>(16.5) | 32 (16) | 65 (16.25) | 6 (6) |  |
|  | **Hereditary** | 12 (6) | 17 (8.5) | 47 (11.75) | 0 (0) |  |
|  | **Impure blood** | 11 (5.5) | 14 (7) | 33 (8.25) | 1 (1) |  |
|  | **Other: God's will, karma, impure blood, witchcraft, immoral conduct, punishment for sins** | 11 (5.5) | 17 (8.5) | 58 (14.5) | 4 (4) |  |
| **Transmission of Leprosy** | **Don't know** | 30 (15) | 22 (11) | 48 (12) | 7 (7) |  |
|  | **Skin contact** | 66 (33) | 80 (40) | 171 (42.75) | 20 (20) |  |
|  | **Eating together** | 49<br>(24.5) | 54 (27) | 111 (27.75) | 8 (8) |  |
|  | **Other:**<br>**contaminated**<br>**soil, sexual**<br>**contact,**<br>**shaking hands,**<br>**sharing**<br>**personal items,**<br>**'different** | 26 (13) | 38 (19) | 103 (25.75) | 6 (6) |  |
|  | **By air** | 122 (61) | 118 (59) | 205 (51.25) | 86 (86) | 35% |
| **Treatability**<br>**of Leprosy** | **Can be treated** | 197<br>(98.5) | 196 (98) | 387 (96.75) | 98 (98) | 92% |
|  | **Don't know** | 3 (1.5) | 2 (1) | 11 (2.75) | 2 (2) |  |
|  | **Can't be**<br>**treated** | 9 (4.5) | 8 (4) | 18 (4.5) | 0 (0) |  |
|  | **Avoiding taboo**<br>**food, herbs,**<br>**rituals,**<br>**differently** | 2 (1) | 5 (2.5) | 8 (2) | 1 (1) |  |
| **Treated how** | **By medication** | 197<br>(98.5) | 196 (98) | 387 (96.75) | 98 (98) | 92% |
|  | **Other** | 14 (7) | 15 (7.5) | 37 (9.25) | 3 (3) |  |
| **Disabilities** | **Disabilities can be prevented** | 161<br>(80.5) | 155<br>(77.5) | 302 (75.5) | 89 (89) | 79% |
|  | **Don't know** | 19 (9.5) | 16 (8) | 33 (8.25) | 5 (5) |  |
|  | **Disabilities can't be prevented** | 20 (10) | 29 (14.5) | 65 (16.25) | 6 (6) |  |
| **Duration of condition** | **Leprosy is temporary** | 197<br>(98.5) | 198 (99) | 383 (95.75) | 99 (99) | 97% |
|  | **Leprosy is permanent** | 1 (0.5) | 0 (0) | 6 (1.5) | 0 (0) |  |
|  | **Don't know** | 2 (1) | 2 (1) | 11 (2.75) | 1 (1) |  |

### Questions for index patients only

The final five questions of the KAP questionnaire were asked to index patients (*n=*200) only. Responses from persons affected by leprosy reveal that while the majority reported no self-withdrawal or community discrimination, a notable proportion still experience residual stigma. About 42.5% preferred to conceal their disease status, indicating persistent fear of disclosure. Conversely, most respondents disagreed that they were disrespected (71.5%) or socially excluded (81%) due to leprosy. Only 4% admitted avoiding social or work interactions by choice. These findings suggest that enacted stigma has substantially declined, yet internalized stigma remains evident among a subset of patients, emphasizing the need for continued psychosocial support and community awareness (Fig 1.)

**Fig 1.**
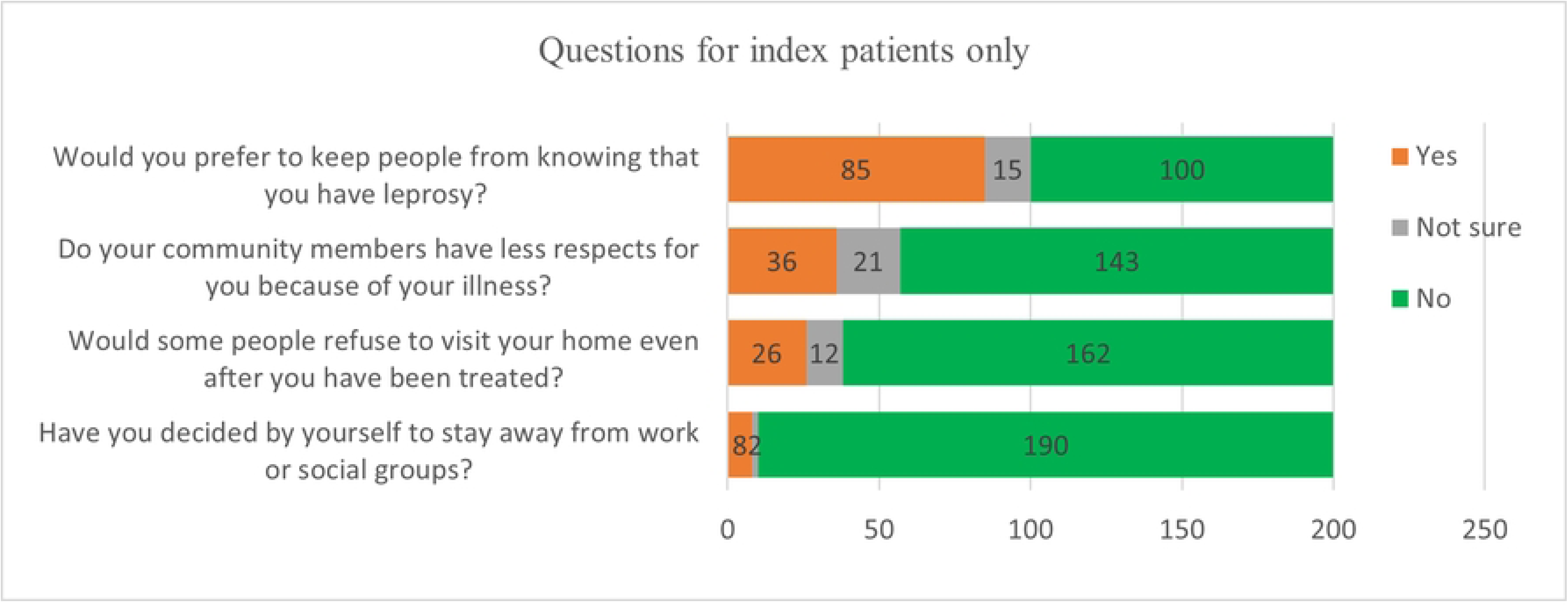
Distribution of participant responses to the KAP questions.

### Overall knowledge assessment among all groups

Analysis of knowledge levels across different participant groups revealed significant disparities in awareness and understanding of leprosy-related concepts. Among persons affected by leprosy, the majority demonstrated adequate knowledge, with 69% scoring seven or more out of nine, while only 4% showed poor knowledge. One of the leprosy index patients mentioned during in-depth interview that

> *“This is a communicable disease which happen by coming close to the leprosy patient or it occurs through touching” [**In-depth interview; index patient, Female, 36-40 Y**]*

A similar pattern was observed among contacts, where 67% had adequate knowledge and just 2% fell in the poor category.

Knowledge levels were comparatively lower in the general community group, where 56% showed adequate knowledge and 6% demonstrated poor understanding, indicating a gap in knowledge dissemination at the community level. Health workers exhibited the highest level of awareness, with 89% achieving adequate knowledge scores and only 1% showing poor knowledge. This underscores the effectiveness of formal training and professional exposure among health workers.

Overall, the data indicate that persons affected by leprosy and their close contacts possess relatively high levels of disease-specific knowledge, whereas the general community lags behind. The significant difference across groups highlights the need for broader community-based educational interventions to complement existing programs targeted at patients and health professionals. This is shown in the table 3.

**Table 3:** Level of Knowledge among all participants group.

| Participant type | Responses were given as a percentage of participants who gave the correct answer even if incorrect answers were present *n* (%). |  |  |
| --- | --- | --- | --- |
|  | Poor knowledge | Moderate knowledge | Adequate knowledge |
|  | Three or less out of nine | Score between four and six | Score of seven or more |
| **Persons affected (n = 200)** | 7 (4) | 55 (28) | 138 (69) |
| **Contacts (n = 200)** | 3 (2) | 64 (32) | 133 (67) |
| **Community (n = 400)** | 24 (6) | 153 (38) | 223 (56) |
| **Health workers (n = 100)** | 1 (1) | 10 (10) | 89 (89) |

### Relationship between sociodemographic characteristics and KAP score

All the participants were assessed to determine the predictors of leprosy-related knowledge, attitudes, and practices (KAP) across three distinct participant groups: index patients, contacts/community members, and health workers. The findings revealed significant sociodemographic and clinical determinants influencing KAP within and across these groups.

Among index patients, education level (p = 0.002) and disability grade (p = 0.046) showed significant associations with KAP levels. Regression analysis further identified that multibacillary (MB) type (β = 1.14, p < 0.001) and higher education were strong predictors of favorable KAP, while Grade 2 disability and increasing age negatively influenced KAP.

For contacts and community members, higher education (p = 0.001), marital status (p = 0.001), and non-Islamic religion (p = 0.023) were associated with better KAP scores. Regression modeling revealed that female gender (β = 0.382, p = 0.015), higher education (p < 0.003), marital status (β = 0.713, p = 0.006), and non-Islamic faith (β = 0.682, p = 0.001) were independent predictors of higher KAP.

Among health workers, education (p = 0.016) and marital status (p = 0.011) were significant. Higher education (β = 0.851, p = 0.028) and being married (β = 2.682, p < 0.001) predicted better KAP, although initial associations with gender and religion were not confirmed in multivariable models.

When all participants were assessed together, education (p < 0.001), marital status (p < 0.001), religion (p = 0.009), area of residence (p = 0.035), and occupation (p = 0.030) emerged as significant factors. Regression analysis confirmed that higher education and being married consistently predicted better KAP (both p < 0.001), while non-Islamic religion also retained significance (β = 0.608, p < 0.001).

Bootstrapped regression models were applied to ensure robustness of results, accounting for non-normal data distribution. Across all groups, higher education and marital status emerged as core determinants of leprosy KAP, underscoring the role of social and educational interventions in enhancing knowledge and promoting early case detection and adherence to treatment. This is shown in the table 4.

**Table 4.** Correlations between level of knowledge about leprosy and the other variables in the dataset.

| **Group** | **Predictor** | **Category** | **Coefficient**<br>**($\beta$)** | **Bootstrap**<br>**SE** | **p-value** | **95% CI** |
| --- | --- | --- | --- | --- | --- | --- |
| **Index**<br>**patient**<br>**(n=200)** | **Age** | Age (years) | -0.0224 | 0.0102 | 0.028 | -0.0423, -0.0025 |
|  | **Leprosy**<br>**type** | PB | Referent | Referent | Referent | Referent |
|  |  | MB | 1.1410 | 0.3024 | <0.001 | 0.5484, 1.7337 |
|  | **Disabilit**<br>**y grade** | 0 | Referent | Referent | Referent | Referent |
|  |  | 1 | 0.8654 | 0.4428 | 0.051 | -0.0025, 1.7333 |
|  |  | 2 | -2.1621 | 0.6691 | 0.001 | -3.4736, -0.8507 |
|  | **Educatio**<br>**n** | Illiterate | Referent | Referent | Referent | Referent |
|  |  | Primary | 0.5721 | 0.2686 | 0.033 | 0.0457, 1.0986 |
|  |  | Secondary | 0.6300 | 0.3322 | 0.058 | -0.0211, 1.2812 |
|  |  | Higher | 1.0598 | 0.5101 | 0.038 | 0.0601, 2.0596 |
|  | **Constant** | - | 7.7002 | 0.5190 | <0.001 | 6.6829, 8.7175 |
| **Contacts/c**<br>**ommunity**<br>**members**<br>**(n=600)** | **Age** | Age (years) | 0.0161 | 0.0064 | 0.012 | 0.0035, 0.0287 |
|  | **Educatio**<br>**n** | Illiterate | Referent | Referent | Referent | Referent |
|  |  | Primary | 0.2667 | 0.1948 | 0.171 | -0.1152, 0.6486 |
|  |  | Secondary | 0.7199 | 0.2412 | 0.003 | 0.2472, 1.1926 |
|  |  | Higher | 1.6056 | 0.2782 | <0.001 | 1.0604, 2.1508 |
|  | **Marital status** | No married | Referent | Referent | Referent | Referent |
|  |  | Married | 0.7128 | 0.2574 | 0.006 | 0.2083, 1.2173 |
|  | **Constant** | — | 4.9097 | 0.3412 | <0.001 | 4.2410, 5.5785 |
| **Health workers (n=100)** | **Educational** | Secondary | Referent | Referent | Referent | Referent |
|  |  | Higher | 0.8506 | 0.3863 | 0.028 | 0.0936, 1.6077 |
|  | **Marital status** | No married | Referent | Referent | Referent | Referent |
|  |  | Married | 2.6818 | 0.3688 | <0.001 | 1.9590, 3.4046 |
|  | **Constant** | — | 5.0000 | - | - | - |
| **All participants (n=900)** | **Educational** | Illiterate | Referent | Referent | Referent | Referent |
|  |  | Primary | 0.2286 | 0.1551 | 0.140 | -0.0754, 0.5326 |
|  |  | Secondary | 0.6632 | 0.1704 | <0.001 | 0.3293, 0.9972 |
|  |  | Higher | 1.6112 | 0.1600 | <0.001 | 1.2976, 1.9248 |
|  | **Marital status** | No married | Referent | Referent | Referent | Referent |
|  |  | Married | 0.9952 | 0.1796 | <0.001 | 0.6431, 1.3473 |
|  | **Constant** | — | 5.6598 | 0.1967 | <0.001 | 5.2743, 6.0454 |

## Discussion

This baseline study among index patients, close contacts, community members and health workers in two high-endemic districts of northwest Bangladesh found generally high levels of leprosy-related knowledge, particularly regarding curability and treatment, alongside important gaps and misconceptions about aetiology and transmission. Overall KAP scores were highest among health workers, followed by persons affected by leprosy and their close contacts, while community members lagged behind. Education, marital status and religion emerged as consistent determinants of more favorable KAP, whereas older age and grade 2 disability among index patients were associated with poorer scores. Qualitative findings suggested that biomedical understanding often co-exists with religious or moral explanatory models and that, although overt discrimination appears limited, anticipated and internalized stigma remain common.

The high proportion of participants recognizing skin patches as an early sign of leprosy and awareness that the disease is curable with multidrug therapy is encouraging and consistent with studies from other endemic settings where knowledge of treatment availability is relatively good [5,6,19]. Awareness that patients on treatment are less infectious and that disability can be prevented also suggests that core programme messages have been partially successful. However, recognition of loss of sensation as a key symptom was substantially lower, particularly among community members and contacts, echoing earlier work which shows that failure to recognize “silent” nerve damage contributes to delayed diagnosis and grade 2 disability [13–16]. Strengthening communication around sensory changes and self-examination is therefore critical for early case detection.

Many participants correctly identified germs or bacteria as the cause of leprosy, yet misconceptions such as heredity, unclean environment and supernatural or moral causes (e.g. God’s punishment, karma) were still frequently mentioned, similar to findings from India, Nepal and Nigeria [20,8,15,17,19]. Knowledge about transmission was also mixed: a considerable proportion mentioned close contact or shared food as routes of spread, and only about half correctly indicated airborne transmission from untreated multibacillary cases [1–3, 18]. Such beliefs may sustain avoidance of persons affected by leprosy and their families despite awareness of curability. These patterns underline that communication strategies must explicitly address co-existing explanatory models and not only provide biomedical facts.

As expected, health workers had the highest proportion of adequate knowledge scores, reflecting formal training and professional exposure, but a minority did not reach the “adequate” threshold, indicating the need to reinforce leprosy content in pre-service and in-service curricula. Persons affected by leprosy and their close contacts also showed predominantly adequate knowledge, likely due to counselling during diagnosis and treatment and targeted activities within national programmes. In contrast, community members had lower knowledge scores, in line with studies from India and Nepal where community misconceptions about transmission and deformity remain widespread despite high awareness that leprosy is curable [21,19, 25–27]. This suggests that patient-focused education should be complemented by broader, community-level engagement in high-burden clusters.

Responses to items on stigma reveal an important tension: most index patients reported little overt exclusion, but many preferred to keep their diagnosis secret. Similar discrepancies between low reported enacted stigma and high internalized or anticipated stigma have been described in leprosy and mental health research [5,23,25–26]. Fear of rejection, shame and concerns about marriage or employment may discourage disclosure and delay care-seeking, even when people understand that leprosy is treatable. Programmes should therefore combine information on curability with strategies that address emotional and social dimensions of stigma, including counselling, peer support and engagement with community and religious leaders [22,25,26,27].

Education and marital status consistently predicted better KAP across groups, echoing findings from other endemic countries where low educational level is associated with poorer knowledge and more stigmatizing attitudes [21,9,19,26]. Health messages should be adapted for people with limited formal education, using simple language, visual materials and interactive methods. Being married may reflect greater social support or more extensive social networks, which can facilitate information exchange. Among index patients, multibacillary disease was associated with higher KAP scores, while older age and grade 2 disability predicted poorer KAP. Patients with more severe disease may have more frequent contact with health services, whereas those with visible disability and older age may have reduced access to information and greater internalized stigma. These findings highlight the need to prioritize older patients and those with grade 2 disability for intensified education and psychosocial support. The association between non-Islamic religion and higher KAP should be interpreted cautiously due to small numbers and may reflect contextual differences rather than religious affiliation.

The study has several strengths, including its relatively large sample size, inclusion of four key stakeholder groups from the same geographic area and use of a mixed-methods design that combines quantitative KAP assessment with qualitative interviews and FGDs. Robust statistical approaches, including non-parametric tests and bootstrapped regression models, were applied to account for non-normal distributions. Limitations include the cross-sectional design, which precludes causal inference, and the potential for social desirability bias, particularly among health workers and community leaders. Stigma and discrimination are sensitive topics and may have been under-reported. The study was conducted in two districts with long-standing leprosy services and ongoing PEP++ activities, so generalizability to areas with lower endemicity or weaker programmes may be limited.

These findings have practical implications for leprosy control and post-exposure prophylaxis. Educational messages should emphasize early symptoms, especially sensory loss, realistic transmission routes and disability prevention, while directly challenging persistent misconceptions. Communication strategies need to be adapted for people with limited education and extended beyond patients to the wider community. For persons affected by leprosy, particularly those with visible disability, psychosocial support and peer-led self-care groups may help reduce internalized stigma and improve health-seeking behavior. Longitudinal studies embedded within PEP++ and similar initiatives could assess how KAP and stigma evolve over time and whether targeted communication strategies contribute to earlier diagnosis, better adherence and reduced transmission.

In summary, this study shows that in two high-endemic districts of Bangladesh, knowledge about the curability and treatment of leprosy is generally high, but important gaps and misconceptions remain, especially among community members. Education and marital status consistently predicted more favorable KAP, while older age and grade 2 disability were associated with poorer scores among index patients. Addressing these knowledge gaps and the more hidden forms of stigma through context-specific, community-based and psychosocial interventions will be essential to support early case detection, effective implementation of post-exposure prophylaxis and progress towards leprosy elimination in Bangladesh and similar settings.

## Limitations

This study has several limitations. First, its cross-sectional design precludes any causal inference or assessment of how knowledge, attitudes and practices evolve over time. Second, all KAP data were based on interviewer-administered, self-reported questionnaires, so social desirability bias is likely, particularly for sensitive items on stigma and discrimination and may have led to under-reporting of negative attitudes or exclusionary behavior. Third, the study was conducted in only two districts of northwest Bangladesh where leprosy services are relatively well established and PEP++ activities are ongoing, which may limit the generalizability of the findings to areas with lower endemicity or weaker programmes. Fourth, although we included four key stakeholder groups, the sample size for health workers was smaller than for community members and may have reduced power for some subgroup analyses. Finally, the KAP thresholds and the “correct-only” scoring approach, while informed by previous leprosy KAP work, may not fully capture the nuances of partial knowledge, ambivalent attitudes or context-specific practices, and the qualitative component was conducted in a limited number of interviews and FGDs, which may not reflect the full diversity of experiences in all communities.

## Conclusion

This baseline mixed-methods study in two high-endemic districts of northwest Bangladesh shows that while core messages on the curability of leprosy, effectiveness of multidrug therapy and prevention of disability have largely reached patients, contacts, communities and health workers, important gaps and misconceptions about aetiology and transmission remain, particularly among community members. Higher education and being married consistently predicted more favorable knowledge, whereas older age and grade 2 disability among index patients were associated with poorer scores. Alongside generally high awareness, internalized stigma persisted among a substantial proportion of persons affected, indicating that fear of disclosure and social consequences continues to influence their lives.

To support early case detection, effective roll-out of post-exposure prophylaxis and progress towards leprosy elimination, leprosy control programs should strengthen context-specific, literacy-sensitive health education that addresses both biomedical facts and prevailing cultural, religious and moral explanatory models. Interventions need to extend beyond patients to their close contacts and the wider community, and should prioritize older individuals and those with visible disability for intensified education and psychosocial support. Integrating structured counselling, peer support and community engagement into routine services, and following these cohorts over time, will be essential to reduce hidden stigma, improve health-seeking behavior and sustain the gains achieved in leprosy control in Bangladesh and similar settings.

## Data Availability

All the data are available in the Management information system of The Leprosy Mission International-Bangladesh. We will provide the dataset if the manuscript accepted for publication or upon the requirement of editorial bodies.

## Acknowledgments

We are grateful to the contributions of all of the participants who shared their knowledge, perceptions, ideas and stories with us. We thank the research team in Nilphamari and rangpur who collected and entered the data and provided logistical support for the study: Md. Anwar Hossain & Md. Rashidul Hasan (district supervisors), Shahidunnahar Reba, Touhidur Rahman Chowdhury, Md. Asaduzzaman Liton, Md. Moshiur Rahman, Mukti Rani Sarker, Baburam Roy (research supervisors) and Ms. Shahana Sultana (research assistant). We are grateful to the support of the principal investigator of the PEP++ project: Wim H. van Brakel, and the international manager of the PEP++ project: Duane Hinders.

## Supporting Information

S1 Checklist. STROBE checklist for reporting the study.

## References

1. Bratschi MW, Steinmann P, Nnawickenden A, Gillis T. Current knowledge on Mycobacterium leprae transmission: a systematic literature review. Lepr Rev. [Internet]. 2015; 86: 142–155. Available from: https://pdfs.semanticscholar.org/2dc5/5a0086c3e3e8a87292552b50c79108 e6e4a1.pdf PMID: 26502685

2. Lastó ria JC, Abreu MAMM de. Leprosy: review of the epidemiological, clinical, and etiopathogenic aspects—part 1. An Bras Dermatol [Internet]. 2014; 89(2):205–18. Available from: http://www.ncbi.nlm.nih.gov/pubmed/24770495

3. International Leprosy Association. CC. International journal of leprosy and other mycobacterial diseases. [Internet]. Vol. 30, International Journal of Leprosy. International Leprosy Association; 1962; 10–18 p. Available from: https://www.cabdirect.org/cabdirect/abstract/ 19632900058

4. Control of Neglected Tropical Diseases (NTD). Global leprosy (Hansen disease) update, 2022: new paradigm – control to elimination [Internet]. who.int. 2023; Available from: https://www.who.int/publications/i/item/who-wer9837-409-430

5. Tsutsumi A, Izutsu T, Islam MDA, Amed JU, Nakahara S, Takagi F, et al. Depressive status of leprosy patients in Bangladesh: association with self-perception of stigma. Leprosy Review [Internet]. 2004 Mar 1;75(1):57–66. Available from: 10.47276/lr.75.1.57

6. Joseph GA, Rao PS. Impact of Leprosy on quality of life. Bull World Health Organ 1999;77:515-

7. Da Silva Souza C, Bacha JT. Delayed diagnosis of leprosy and the potential role of educational activities in Brazil. Lepr Rev 2003; 74:249–58. PMID: 14577470

8. Opala J, Boillot F. Leprosy among the Limba: illness and healing in the context of world view. Soc Sci Med 1996; 42:3–19. PMID: 8745104

9. John AS, Rao OS. Awareness and attitudes towards leprosy in urban slums of Kolkata, India. Indian J Lepr 2009; 81:135–40. PMID: 20509342

10. Pickens J. Attitudes and perceptions. Organ Behav Heal Care 2005;4.

11. Hiebert PG. Transforming worldviews: An anthropological understanding of how people change. Baker Academic; 2008.

12. Broadbent E, Wilkes C, Koschwanez H, Weinman J, Norton S, Petrie KJ. A systematic review and meta-analysis of the Brief Illness Perception Questionnaire. Psychol Health 2015; 30:1361–85. 10.1080/08870446.2015.1070851 PMID: 26181764

13. Bekri W, Gebre S, Mengiste A, Saunderson PR, Zewge S. Delay in presentation and start of treatment in leprosy patients: a case-control study of disabled and non-disabled patients in three different settings in Ethiopia. Int J Lepr Other Mycobact Dis 1998; 66:1. PMID: 9614833

14. Nicholls PG, Chhina N, Bro AK, Barkataki P, Kumar R, Withington SG, et al. Factors contributing to delay in diagnosis and start of treatment of leprosy: Analysis of help-seeking narratives in northern Bangladesh and in West Bengal, India. Lepr Rev 2005; 76:35–47. PMID: 15881034

15. Reddy NB, Satpathy SK, Krishnan SA, Srinivasan T. Social aspects of leprosy: a case study in Zaria, northern Nigeria. Lepr Rev 1985; 56:23. PMID: 3990504

16. Robertson LM, Nicholls PG, Butlin R. Delay in presentation and start of treatment in leprosy: experience in an out-patient clinic in Nepal. Lepr Rev 2000; 71:511–6. PMID: 11201907

17. Van de Weg N, Post EB, Lucassen R, De Jong J, Van Den Broek J. Explanatory models and help-seeking behaviour of leprosy patients in Adamawa State, Nigeria. Lepr Rev 1998; 69:382–9. PMID: 9927811

18. Suzuki K, Akama T, Kawashima A, Yoshihara A, Yotsu RR, Ishii N. Current status of leprosy: epidemiology, basic science and clinical perspectives. J Dermatol 2012; 39:121–9. 10.1111/j.1346-8138.2011.01370.x PMID: 21973237

19. Van ‘T Noordende AT, Korfage IJ, Lisam S, Arif MA, Kumar A, Van Brakel WH. The role of perceptions and knowledge of leprosy in the elimination of leprosy: A baseline study in Fatehpur district, northern India. PLoS Neglected Tropical Diseases [Internet]. 2019 Apr 5;13(4): e0007302. Available from: 10.1371/journal.pntd.0007302

20. De Stigter DH, De Geus L, Heynders ML. Leprosy: between acceptance and segregation. Community behaviour towards persons affected by leprosy in eastern Nepal. Lepr Rev [Internet]. 2000 [cited 2018 Sep 11]; 71: 492–498. Available from: https://pdfs.semanticscholar.org/7625/

21. Singh R, Singh B, Mahato S. Community knowledge, attitude, and perceived stigma of leprosy amongst community members living in Dhanusha and Parsa districts of Southern Central Nepal. PLoS Neglected Tropical Diseases [Internet]. 2019 Jan 11;13(1):e0007075. Available from: 10.1371/journal.pntd.0007075

22. Wong ML. Designing Programmes to address stigma in leprosy: Issues and challenges. Asia Pacific Disability Rehabil J 2004;15:3–12.

23. Link BG, Struening EL, Rahav M, Phelan JC, Nuttbrock L. On stigma and its consequences: evidence from a longitudinal study of men with dual diagnoses of mental illness and substance abuse. J Health Soc Behav 1997:177–90. PMID: 9212538

24. Thornicroft G, Rose D, Kassam A, Sartorius N. Stigma: ignorance, prejudice or discrimination? Br J Psychiatry 2007; 190:192–3. 10.1192/bjp.bp.106.025791 PMID: 17329736

25. Weiss MG, Ramakrishna J, Somma D. Health-related stigma: rethinking concepts and interventions. Psychol Health Med 2006; 11:277–87. 10.1080/13548500600595053 PMID: 17130065

26. Ballering AV, Peters RMH, Waltz MM, Arif MA, Mishra CP, Van Brakel WH. Community stigma and desired social distance towards people affected by leprosy in Chandauli District, India. Leprosy Review [Internet]. 2019 Dec 1;90(4):418–32. Available from: 10.47276/lr.90.4.418

27. Mieras L, Singh MK, Manglani PR, Arif MM, Banstola NL, Pandey B, et al. A single dose of rifampicin to prevent leprosy; quantitative analysis of impact on perception, attitudes and behaviour of persons affected, contacts and community members towards leprosy in India, Nepal and Indonesia. Leprosy Review [Internet]. 2020 Dec 1;91(4):314–27. Available from: https://leprosyreview.org/article/91/4/20-20015

